# Stochastic Approximator of Motor Threshold (SAMT) for Transcranial Magnetic Stimulation: Online Software and Its Performance in Clinical Studies

**DOI:** 10.1101/2025.06.03.25328918

**Authors:** Boshuo Wang, Vedarsh U. Shah, Lari M. Koponen, Andrada D. Neacsiu, Zafiris J. Daskalakis, Paul B. Fitzgerald, Lawrence G. Appelbaum, Jessica Y. Choi, Nimesha Gerlus, Yiru Li, Itay Hadas, Yinming Sun, Mohsen Poorganji, Hadley Daniels, Katie Rodriguez, Efstathia Stephanie Gotsis, Neil W. Bailey, Jeydhurga Raveendran, Shona K. Brinley, Alexander T. Gallo, Stefan M. Goetz, Angel V. Peterchev

## Abstract

**Background:** The motor threshold (MT) plays a central role in probing brain excitability and individualizing transcranial magnetic stimulation (TMS). Previously, we proposed stochastic approximation (SA) as a new method for determining TMS MT and demonstrated its excellent speed and accuracy via simulations. SA also has low computational requirements and is robust to potential model flaws.

**Objective:** This project aimed to develop a practical SA thresholding method and assess its performance in clinical studies.

**Methods:** The SA thresholding method was implemented as an online software application—SAMT (Stochastic Approximator of MT)—that incorporated features for detection of inaccurate MT estimation. Two ongoing clinical studies use SAMT and have collected 281 finger muscle MTs from 124 participants to date. SAMT’s misestimation detection method marked MTs of 7 thresholding trials as inaccurate, and SAMT’s performance in the remaining 274 trials was assessed by comparing the MT at each step to the threshold estimated by fitting a sigmoidal probability distribution to the complete muscle response data from the session.

**Results:** By the 25^th^ TMS pulse, 99% of the SAMT MTs deviated by less than 3.0% (relative) and 1.3% of maximum stimulator output (absolute) from the corresponding fitted sigmoid thresholds and were within their 95% confidence intervals.

**Conclusions:** We provide the TMS community with a new motor thresholding tool, SAMT. Combined with the prior simulation results, the experimental assessment presented here supports the practicality and accuracy of the SA thresholding method and the SAMT software.

## Introduction

Transcranial magnetic stimulation (TMS) is a method for activating cortical neurons and modulating neural circuits noninvasively [1,2], with broad research and clinical applications. The motor threshold (MT) of TMS is a critical parameter for cortical excitability measurements, neurological assessment of motor functions, and dose individualization of treatments in non-motor cortical areas. However, TMS thresholding procedures are challenged by the high response variability in cortical neurons and the corticospinal tract [3–5]. Thresholding methods based on relative frequency, including the commonly used five-out-of-ten method, are slow and inherently inaccurate [6–10]. In comparison, adaptive methods of parametric estimation by sequential testing (PEST^1^) using maximum likelihood estimation (MLE) are much faster and accurate [11,16]. However, they rely on an underlying response model that could potentially be inaccurate, biased, or not representative of individual subjects or TMS devices, and currently available implementations require software installation [14,15]. Moreover, these and other methods can result in misestimation under certain conditions, and it is therefore important to incorporate features to check for possible convergence to inaccurate MT estimates [13].

To address these issues, we recently developed a novel class of TMS thresholding methods based on stochastic approximation (SA) [6], a simple algorithm with extremely low computational requirements that is not reliant on an estimation model. Using a computational model of the motor evoked response and a virtual population of 25,000 subjects [17], we demonstrated that SA methods are comparable or superior to existing adaptive PEST methods in terms of accuracy and speed [6].

To enable the TMS community to use the SA methods, we developed an online application SAMT (Stochastic Approximator of Motor Threshold, [18]). SAMT includes instructions for selecting the initial pulse amplitude of the thresholding procedure to mitigate inaccurate estimations and a heuristic for detecting most inaccurate estimates at the end of the procedure [13]. In this study, we describe the details of SAMT and assess its performance in two ongoing clinical studies.

## Methods

### Overview of stochastic approximation for TMS thresholding

As SA methods [19,20] were only recently introduced to TMS thresholding procedures [6], we briefly summarize their key steps and characteristics. SA generates a series of TMS stimulation amplitudes, *x*_*i*_, using a sequence that statistically converges towards the threshold, *x*_*t*_:

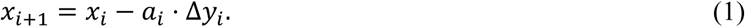

Here, *i* ≥ 1 is the pulse number, Δ*y*_*i*_ is the deviation of the TMS response from the target level, and *a*_*i*_ is the step size of the sequence. Similar to parametric estimation methods, the stimulation amplitudes outputted by SA methods provide the iteratively updated threshold estimate, *x*_*i*+1_, which is both the threshold estimate at the current pulse and the stimulation amplitude at the next pulse.

We previously explored several options for the control sequence (analog or digital, and fixed or adaptive stepping) and the convergence of the step size *a*_*i*_ (harmonic, geometric, or mixed) [6]. Unlike the analog control sequence that utilizes the logarithmic difference between the amplitude of the motor evoked potentials (MEPs) from electromyography (EMG) recordings and a reference level (typically 50 μV) to calculate Δ*y*_*i*_, the digital control sequence (DCS) uses a binary sequence Δ*y*_*i*_ = ±1 to denote supra- and sub-threshold responses. Thus, DCS SA methods are more convenient as they can be used by observing muscle responses either visually or via EMG; they also appear more accurate without the intrinsic bias of analog sequences [6]. In fixed stepping, the step size follows a predetermined sequence, e.g., *a*_*i*_ = *a*_0_/*i* for harmonic convergence, with *a*_0_ being the initial step size. Adaptative stepping, in contrast, only adjusts the step size following a sign change in the control sequence Δ*y*_*i*_, i.e., when the response changes from subthreshold to suprathreshold or vice versa from the previous step. This feature accelerates the convergence by approaching the threshold faster with a slower decreasing step size when the stimulation amplitude is far from threshold and allows the procedure to consistently reach perithreshold TMS pulse amplitudes. In a population of 25,000 virtual subjects, we showed that the best-performing SA methods achieved a median relative threshold error smaller than relative frequency methods within 20 steps and, depending on the variant, had either similar or smaller errors than adaptive PEST [6].

### TMS motor thresholding tool SAMT

SAMT (current version 1.8.0, example screenshot shown in Figure 1) is implemented in HTML/JavaScript and hosted on GitHub (https://tms-samt.github.io, [18]) since April 2023. It can be accessed via any web browser on any platform or device and does not require download or software installation, in contrast to existing parametric estimation tools [14,15]. The application can also be stored locally as an HTML file and run in a web browser without internet access. SAMT is offered under (a) an open-source license under the GPLv2 license for non-commercial use, or (b) a custom license with Duke University, for commercial use without the GPLv2 license restrictions. SAMT’s layout dynamically adapts to the screen width, with a vertical layout suitable for mobile phones, and a wider one with contents spread out horizontally for computers and tablets. SAMT’s processing is performed locally by the browser and no data from the thresholding procedure are transmitted or stored online, ensuring privacy. The users’ choices for the settings are remembered locally by the browser. Keyboard shortcuts are provided as a fast and convenient alternative method of some operations, such as starting or resetting the procedure, inputting the response, and downloading or copying a record of the procedure.

**Figure 1:**
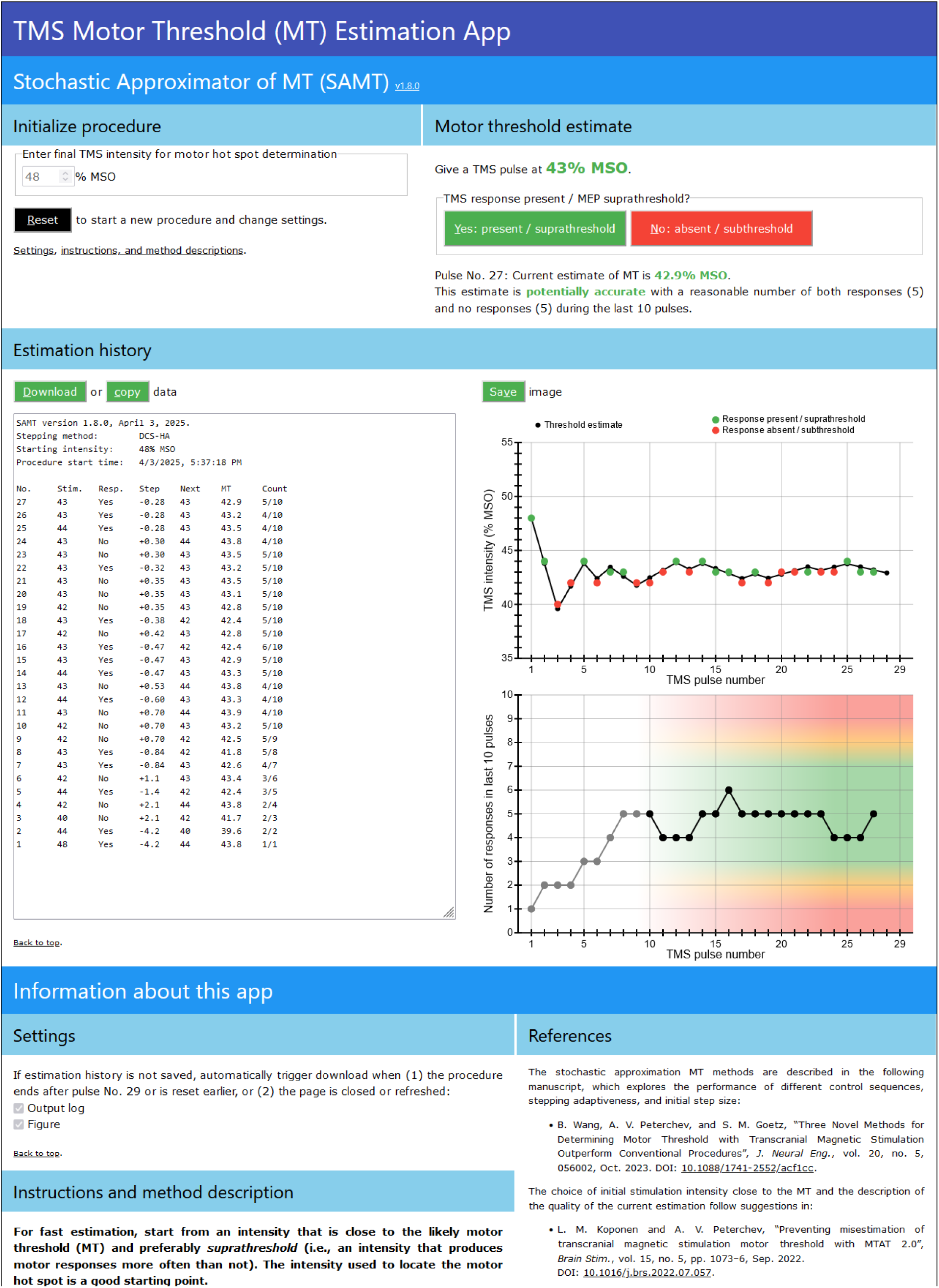
SAMT user interface. Screenshot of SAMT (version 1.8.0, horizontal layout) in use, after 27 pulses with the initial amplitude set to 48% MSO.

SAMT implements SA methods using DCS with the step size following harmonic convergence and a fixed (DCS-H, available up to version 1.7.4) or adaptive (DCS-HA, included since version 1.6.0) stepping sequence. The initial step sizes *a*_0_ in SAMT uses the optimal value previously determined, 6.7% and 4.2% maximum stimulator output (MSO) for fixed and adaptive stepping, respectively [6]. DCS-H was initially selected as it had the fastest convergence and smallest errors among all SA methods previously explored and even outperformed PEST as well as other candidate methods [6]. However, this conclusion was potentially specific to the simulation testbed and testing conditions—including the MEP response model [21,22,17] and the suprathreshold starting amplitudes with a minimum 95% response probability for each virtual subject—and thus may not be generalizable. Therefore, the adaptive DCS-HA was later included^2^, which had performance similar to DCS-H and could provide faster and more robust convergence for other cases not represented in our previous work, e.g., when the starting amplitude is far from the MT, TMS is applied with non-conventional devices and/or pulse shapes [23–26], or the targeted muscle has an input–output curve with slopes different from that of the hand muscles [27] represented in the MEP model.

SAMT currently incorporates two features to mitigate the possibility of misestimations. Firstly, it instructs the user to start the procedure with the final TMS amplitude used for motor hot-spot hunting, which is typically in the vicinity of the MT [13]. Secondly, SAMT counts the responses over the last 10 pulses during the procedure to detect and warn the user against potential inaccurate estimates [13]. Like PEST, the SA methods use the threshold estimate as the next pulse amplitude and therefore the response rate should be approximately 50% as the sequence converges on the MT with decreasing step size *a*_*i*_. For stimulation amplitudes near the threshold, the procedure is very unlikely to have extreme cases where the last 10 pulses result in 0, 1, 9, or 10 responses. More specifically, given a 50% response rate, a sequence of 10 pulses should have a 0.98% probability of having only one response according to the binomial distribution. Conversely, the likelihood of observing a one-out-of-ten outcome is much higher for a subthreshold response rate of less than 50% than the likelihood for a response rate of 50%. Therefore, considering the observation of a single response during the last 10 pulses as a misestimation will result in at most 1% of accurate estimates (or one out of 102 trials^3^) being falsely rejected. The situation is even more extreme (< 0.1%) for observing no responses, and the same conclusions apply for observing 9 or 10 responses, respectively. Therefore, SAMT marks these estimates as likely inaccurate. SAMT further cautions the users when the last 10 pulses result in 2 or 8 responses. While directly considering these two borderline cases inaccurate would result in too many accurate estimates being falsely rejected (4.4%, or about one out of 23 trials), the users are advised to evaluate the thresholding procedure before accepting the estimate. For example, if eight responses occur consecutively at the end of the sequence of the last 10 pulses, the estimate at the end could be inaccurate, which SAMT warns the user about.

A thresholding trial with SAMT starts with the initial amplitude entered by the user. With each TMS pulse, the user enters whether a suprathreshold response is evoked or not via mouse clicks or keyboard shortcuts. Depending on the method of detection, a suprathreshold response can be, for example, a peak-to-peak MEP amplitude ≥ 50 µV or a visible muscle twitch. SAMT then provides the threshold estimate (rounded to 0.1% MSO for display) and next stimulation amplitude (rounded to 1% MSO). Both the estimate and next amplitude are clamped when the underlying SA procedure attempts to step below 1% MSO or above 100% MSO, although the user should still be cautious if such a situation is encountered unless a very high or a very low MT is expected based on the TMS device configuration and the hot-spot hunt. The information of the procedure is displayed, with the TMS pulse amplitudes, responses, threshold estimates, and additional data tabulated in reverse order of the pulse number. Two graphs provide visualization of the procedure by plotting the stimulation amplitude pulse by pulse and the number of responses during the last 10 pulses, which further help the users evaluate the accuracy of the MT estimate. After 20 pulses, when the median relative error in our modeling study was less than 1.5% [6], SAMT provides a description of the accuracy of the MT using the response count from earlier pulses^4^. The user may continue until 29 pulses to improve the accuracy, at which point SAMT automatically stops. If the threshold estimate is considered unlikely to be accurate, SAMT prompts the user to choose a new initial amplitude either higher or lower than the previous one depending on the number of responses during the last 10 pulses and restart the trial. Alternatively, the user can refine the threshold estimate from the last pulse amplitude of the current trial, for which SAMT automatically transfers this information to restart a new thresholding trial.

### SAMT in clinical studies

Utilizing SAMT, TMS MTs were measured in two ongoing clinical studies, the NATURE study at Duke University (NCT05712057 [28]) and the FREED study at the University of California, San Diego (UCSD) and the Australian National University (ANU) (NCT06003309 [29]). The studies were approved by their respective institutional ethics committees and conformed to the Declaration of Helsinki. Relevant details for the thresholding procedures of the clinical studies are shown in Table 1. Briefly, the MT measurements were performed on study participants using MagPro X100 or R30 stimulators and Cool-B65 or C-B60 coils (MagVenture A/S, Farum, Denmark) with biphasic pulses and the initial induced electric field in the anterior–posterior direction as part of the thresholding before the treatment protocols or assessment pre- and post-treatment. MT was obtained from the left first dorsal interosseous (FDI) in the NATURE study and the right abductor pollicis brevis (APB) in the FREED study. The SAMT output logs (text files) were read with MATLAB (version 2022b, The MathWorks, Natick, MA, USA), which was also used for subsequent data analyses and visualization.

**Table 1.** Relevant information of SAMT application in clinical studies.

| Study | NATURE | FREED |  |
| --- | --- | --- | --- |
| Site | Duke | UCSD | ANU |
| TMS device | MagPro X100<sup>a</sup> | MagPro X100<sup>a</sup> | MagPro R30<sup>a</sup> |
| TMS coil | Cool-B65 A/P<sup>a</sup> | C-B60<sup>a</sup> | Cool-B65<sup>a</sup> |
| Pulse shape | Biphasic | Biphasic | Biphasic |
| Initial electric field direction | Anterior–posterior | Anterior–posterior | Anterior–posterior |
| Muscle target | First dorsal interosseous | Abductor pollicis brevis | Abductor pollicis brevis |
| Muscle side | Left | Right | Right |
| No. of participants | 63 | 33 | 28 |
| Participant sex (female/male) | 52 / 11 | 19 / 14 | 23 / 5 |
| Participant age (range, mean ± SD) | 18–55, 33.8 ± 10.4 | 22–79, 45.1 ± 15.7 | 19–75, 39.5 ± 15.0 |
| Participant handedness (right/left) | 58<sup>b</sup> / 5 | 24 / 7<sup>c</sup> | 27 / 1 |
| No. of SAMT trials | 65 | 115 | 99 |
| Trials per participant (mean ± SD) | 1.0 ± 0.2 | 3.6 ± 0.8 | 3.5 ± 1.2 |
| Stepping (fixed/adaptive) | 43 / 22 | 86 / 29 | 87 / 12 |
| SAMT versions | 1.4.1–1.4.3, 1.4.5, 1.6.2,<br>1.7.1–1.7.5, 1.8.0<sup>d</sup> | 1.4.5, 1.7.0–1.7.5, 1.8.0 | 1.6.0–1.6.1, 1.6.6, 1.7.0–<br>1.7.4, 1.8.0 |

<sup>a</sup> MagVenture A/S, Farum, Denmark
<sup>b</sup> Includes two participants self-identified as ambidextrous.
<sup>c</sup> Handedness not collected for two participants.
<sup>d</sup> Versions 1.3.5 and 1.4.0 used for pilot testing and training but not for data collection.
SD: standard deviation.

A total of 281 thresholding trials were completed on 124 participants between the studies’ respective starting dates and May 1, 2025 (Figure 2; NATURE: 65; FREED: 216 (UCSD: 117; ANU: 99)), with the measurement repeated for some participants for reliability, for comparison pre- and post-TMS treatment, or due to initial inaccurate estimation. SAMT ran for a minimum of 25 pulses for all trials, the minimum number at which SAMT (most versions used in the study) provided an accuracy description for the MT, and 238 trials terminated at the 29^th^ pulse (NATURE: 35; FREED: 203 (UCSD: 104; ANU: 99)), the maximum number at which SAMT automatically stops (Figure S3A). Most trials (218) used fixed stepping, and a small portion of later trials (63) used adaptive stepping. Seven trials (NATURE: 2; FREED: 5 (UCSD: 3; ANU: 2); all fixed stepping and ending at pulse number 29) had either too few (0 or 1) or too many (9 or 10) responses during the last 10 pulses, and were therefore marked as inaccurate estimations by SAMT (Figure 2, squares with red outline; Figure S3A, red lines) according to the criteria for thresholding methods which use the threshold estimate as the next pulse amplitude [13]. Statistics were therefore calculated for the remaining 274 trials, which included 18 trials (NATURE: 1; FREED: 17 (UCSD: 10; ANU: 7); with 2 trials from the UCSD site using adaptive stepping) that were considered borderline accurate. Although excluded, the 7 misestimation trials were analyzed and visualized in the results for comparison (data shown in red).

**Figure 2:**
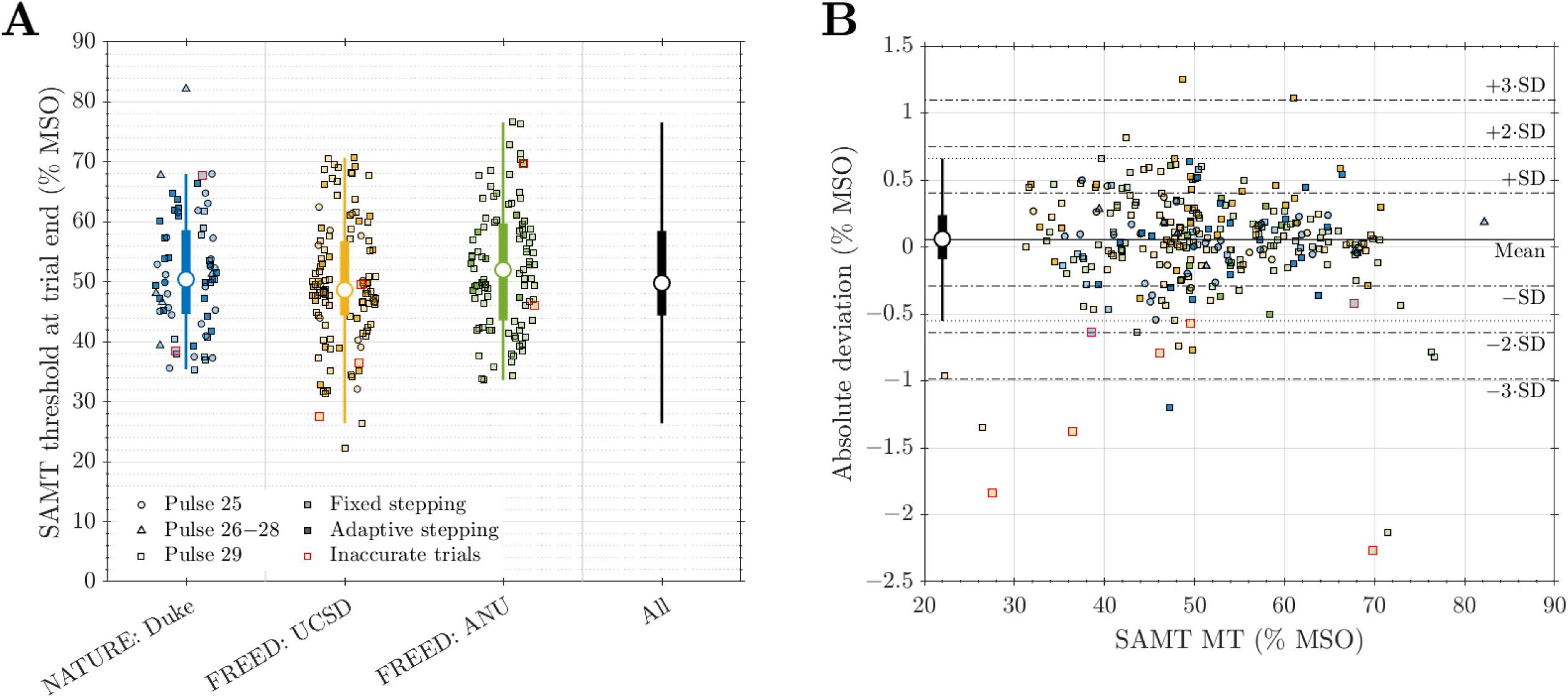
SAMT MTs and their deviations against fitted sigmoid thresholds at trial end. **A.** SAMT MTs at the end of the trials, with the scattered data points showing all 234 individual trials. Trials ending at pulse number 25 (37 trials), between pulse number 26 and number 28 (6 trials), and at pulse number 29 (231 trials) are shown with circle, triangle, and square markers, respectively; estimates obtained with fixed and adaptive stepping have markers with light and dark shading respectively, and those considered inaccurate by SAMT are distinguished with red outlines. Excluding the inaccurate data points, statistics are given for each study site (NATURE: blue; UCSD: yellow; ANU: green), as well as all data (black): box-and-whisker plots show the median (marker), first and third quartiles (box), and lower and upper adjacent values (whisker), which are within 1.5 times the interquartile range below and above the first and third quartiles, respectively. **B**. A modified Bland-Altman plot shows the distribution of absolute threshold deviations at the trial end versus the SAMT threshold estimates, with additional statistics provided by the box-and-whisker and horizontal lines. The marker shape and color represent the same information as in panel A.

### Comparing SAMT thresholds against maximum likelihood estimates

To analyze the accuracy of the MTs obtained with SAMT, we fitted a sigmoidal response probability distribution to the complete sequences of TMS pulses and responses of SAMT trials and estimated the underlying threshold parameter of the sigmoidal distribution, i.e., the amplitude at which the response rate was 50%. Among possible fitting methods such as logistic regression, we used MLE to fit the motor response probability that was described by a cumulative Gaussian distribution function [6,11] with two parameters, the threshold *t* and the spread *s*:

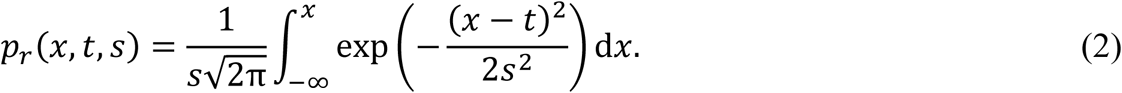

For the purpose of fitting, the response model (2) was simplified by using only the threshold parameter *t*, whereas the spread parameter was fixed to the threshold by a constant ratio based on empirical relationship [6,11]: *s* = 0.07 ⋅*t*. Such simplification was not only for improving the computation speed, but also necessary as a full estimation of two parameters had two major challenges: a much slower convergence due the exploration of the additional dimension in the parameter space [6], and the high inaccuracy and sensitivity in estimating the spread parameter, due to most of the stimulation pulses being applied near the threshold and not able to cover a sufficiently wide range of the response probability curve (2).

The likelihood function for a sequence of stimuli and responses was defined for independent events

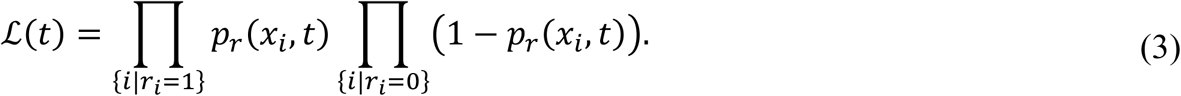

Here, *r*_*i*_ is the binary response of pulse *i*, with *r*_*i*_ = 1 for a response and *r*_*i*_ = 0 for no response. MLE finds *t̂* that maximizes ℒ(*t*), *t̂* = argmax ℒ(*t*), which was equivalently performed on the log-likelihood for numerical robustness and faster calculation. Following [11], two *pseudo-responses* were included in the sequence for robust estimation: one *no response* at 50% MSO below the minimum pulse amplitude of the SAMT sequence (or 0% MSO, if the former is lower) and one *response* at 50% MSO above the maximum of the sequence (not capped at 100% MSO, as higher amplitudes can be theoretically achieved, e.g., by raising the limit of the stimulator or using a different device). It should be emphasized that we applied MLE to fit a sigmoidal probability distribution to the SAMT sequences *post-hoc*, with the pulse amplitudes in the sequence following the stepping of SA methods; this is in contrast to adaptive PEST, which uses the MLE-fitted threshold parameter at each step to determine the next pulse amplitude [6,11].

To compare the MTs at the end of the SAMT trial (*x*_*t*_) against the fitted sigmoid thresholds (*t̂*), analyses followed previous literature [30–32] and included Kolmogorov-Smirnov test for normality and outlier analysis for each dataset, F-test of equality of variances, two-tailed paired *t*-test (equivalently one-sample test on the absolute deviation Δ_*t*_ = *x*_*t*_ − *t̂*), correlation analyses using Pearson correlations and intraclass correlation coefficients (ICC) [33], and Bland-Altman plots for limits of agreement [34]. For all analyses, the significance level was set to *α* = 0.01. To examine the progression of SAMT’s accuracy, the absolute and relative deviations of the MT estimate at pulse *i*, *x*_*t*,*i*_ = *x*_*i*+1_, versus the fitted threshold *t̂* of the entire SAMT procedure (*post-hoc* deviations) were calculated as

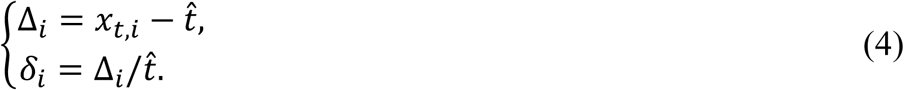

We also performed the parameter fitting with the partial data consisting of stimuli–response pairs from the beginning of the thresholding procedure to a given pulse number *i* (with the same pseudo-responses also included for robust estimation). This estimator, *t̂*_*i*_, provides a running pulse-by-pulse comparison without information of how the procedure would continue. The pulse-by-pulse deviations of *x*_*t*,*i*_ compared to *t̂*_*i*_ are denoted as Δ^_*i*_ and *δ*^_*i*_, and should differ from their *post-hoc* counterparts except at the last pulse.

For each set of partial data, we further calculated the 95% confidence interval (CI) of *t̂*_*i*_, CI_*i*_, using the likelihood-ratio method [35–37]. Briefly, the method compared the likelihood as a function of the parameter to its maximum:

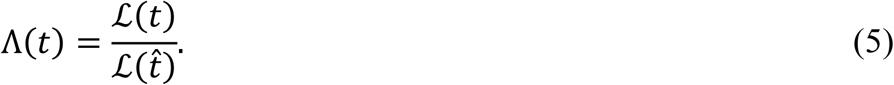

The deviance, defined as twice the negative logarithm of Λ(*t*), followed approximately a chi-square (Χ^2^) distribution with one degree of freedom [35–37]

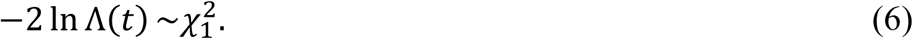

Therefore, the 95% CI was constructed by finding the range of parameter *t* for which the likelihood function satisfied

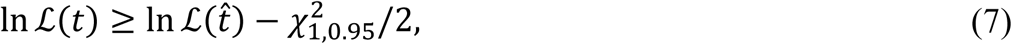

where 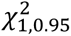 was the 95^th^ percentile of the cumulative distribution function of the 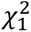 distribution. With 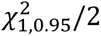 ≈ 1.92, the CI corresponded to the range where the log-likelihood was within approximately two log-units below its maximum value. It should be noted that, depending on the shape of the log-likelihood ln ℒ(*t*), this interval may not necessarily be symmetric around the estimated parameter *t̂*.

## Results

### Motor thresholds

The distribution of SAMT MTs at the trial end are shown in Figure 2A, with numerical values and additional statistics given in Table S1. The MTs had a normal distribution and were in the range between 22.3% and 82.2% MSO, with a median, mean, and standard deviation (SD) of 49.8%, 51.2%, and 10.1% MSO, respectively. Comparing the MTs of the three study sites indicated no statistically significant difference (one-way ANOVA, *p* = 0.17). For repeated measurements on the same participant, the SAMT MTs had trial-by-trial variation (Figure S1), which was overall consistent with test-retest reliability reported for TMS [31,38–41]. Some of the variation can be attributed to different TMS operators and subject variability in pre- versus post-study sessions (FREED study).

### Accuracy of SAMT thresholds

The fitted sigmoid thresholds obtained using the entire SAMT thresholding procedure had similar distribution and statistics as the SAMT MTs (Figure S2A, Table S1), and the variance of the fitted thresholds was equal to that of the SAMT MTs (F = 0.995, *p* = 0.97). The SAMT MTs agreed exceedingly well with the fitted thresholds, showing strong correlation (Pearson *r* = 0.9994, *p* ≪ 0.01, Figure S2B; ICC(A,1) = 0.9994, *p* ≪ 0.01). The absolute deviations at the trial end ranged between −2.14% MSO and 1.25% MSO, had a median, mean, and SD of 0.06% MSO, 0.06% MSO and 0.35% MSO, respectively, with statistically-significant difference from zero (*p* = 0.0098), and were uncorrelated with the thresholds (*r* = −0.060, *p* = 0.32) (Figure 2B, Table S1). The corresponding statistics for relative deviations were respectively [−4.86%, 2.64%], 0.12%, 0.11%±0.77% (*p* = 0.015), and *r* = −0.018, *p* = 0.77 (Figure S2C, Table S1). As the absolute deviations were on average within the 0.1% MSO precision of SAMT’s MT, their statistically-significant difference from zero—an effect dominated by the UCSD site (Table S1)—did not have practical implications on the accuracy. The relative deviations were not significantly different from zero and all within the 5% limit for acceptable estimates [42]. The absolute deviations tended to have higher variance at lower MTs (Fig. 2B, although not statistically significant, Breusch–Pagan test [43] for heteroskedasticity *p* = 0.86), whereas the effect was significant for relative deviations (Fig. S2C, *p* = 3.7×10^−5^). The most likely contributor was the coarser relative changes due to rounding of the stimulation amplitudes (to 1% MSO) and threshold estimates (to 0.1% MSO) in SAMT.

Comparing the MT estimates at each step (Figure S3A) against the fitted threshold at trial end, the absolute deviations of SAMT MTs converged with increasing number of pulses (Figure 3) and reached a median absolute value (thick black line) of less than 1% MSO within 10 pulses. At pulse number 25, the median absolute value was 0.34% MSO and all deviations except for one (3.1% MSO, thin green lines) were within 1.8% MSO, with 99% (271 out of 274 trials) being within 1.3% MSO. Similarly, the relative deviation (Figure S4) had a median absolute value of less than 2% at pulse number 10 and 0.64% at pulse number 25, where all relative deviations were within 3.0% except for the same outlier trial (4.3%). The statistics of the deviations for trials reaching pulse number 29 were further reduced (median 0.18% MSO or 0.37%), with only 5 trials having deviations higher than 1% MSO or 2%. Although it should be noted that the deviations of an individual trial do not necessarily decrease monotonically as the trial proceeds and might fluctuate or slightly increase. Closer inspection reveals that the outlier trial was considered borderline inaccurate by SAMT with two responses during the last 10 pulses and it could have been ruled out by the TMS operator based on the response pattern, i.e., only four responses for the total 29 pulses and two responses during the last 18 pulses (Figure S7). As expected, the pulse-by-pulse deviations (Figure S5), which were calculated against the fitted threshold at the same pulse number (Figure S3B), were smaller than the corresponding *post-hoc* deviations, except at the last pulse for each trial where they were equal. For most trials, the absolute and relative pulse-by-pulse deviations reached less than and stayed respectively within 1% MSO or 2% after only a few (typically between 3 and 5) pulses. Such early agreement of the SAMT MTs to the pulse-by-pulse fitted thresholds demonstrated the very quick convergence speed of SA methods with appropriate starting amplitudes.

**Figure 3:**
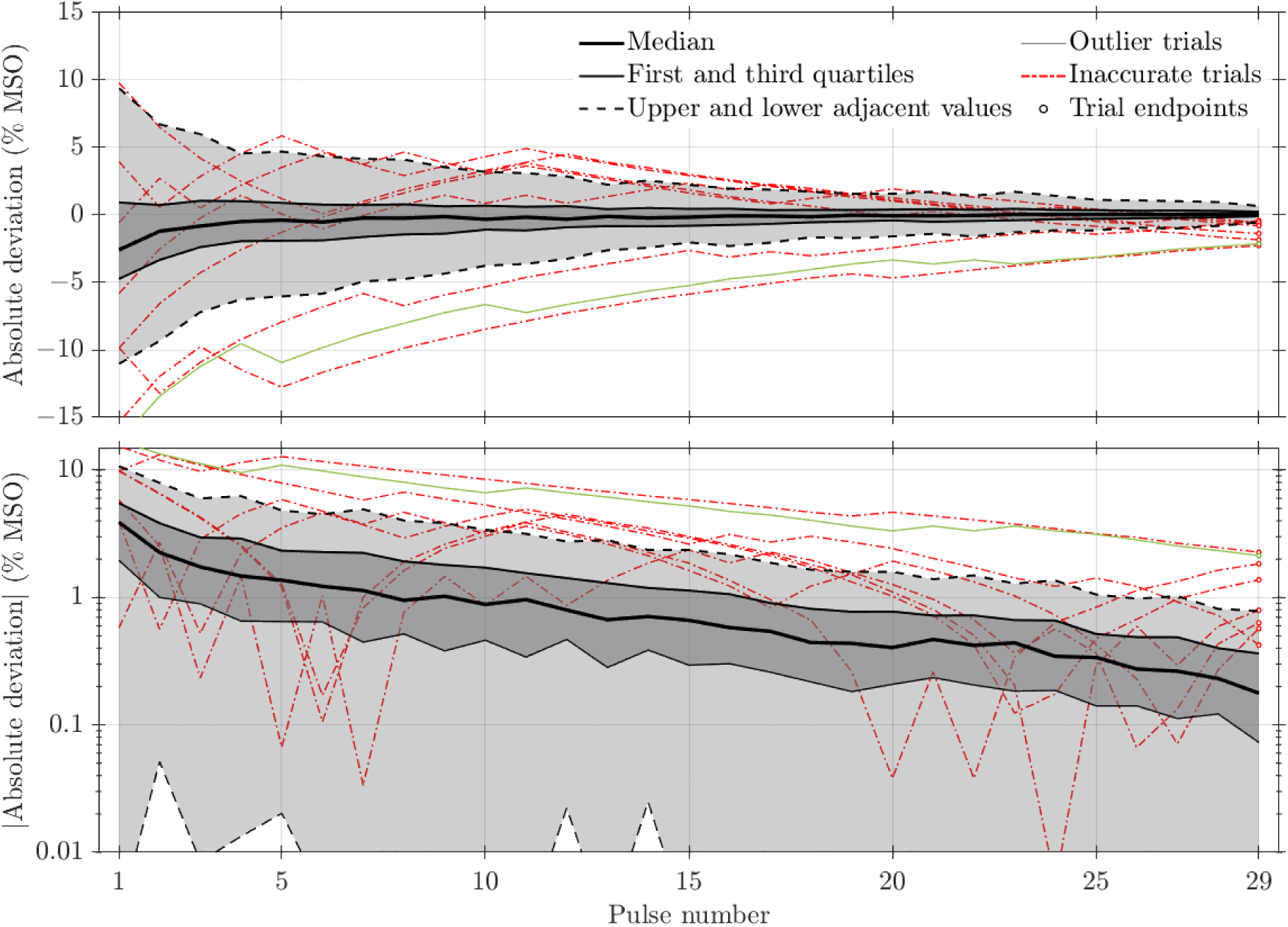
Absolute deviation of SAMT MTs versus number of pulses. Statistics at each pulse number for all trials (274 up to pulse number 25 and fewer trials afterward) are given by black lines, showing median (thick line), first and third quartiles (thin lines with dark gray shading), and lower and upper adjacent values (dashed lines with light gray shading), for the raw deviations (top) and the absolute value of deviations (bottom, on logarithmic scale). The green thin line in each panel represents one individual trial whose deviations were consistently outliers (i.e., outside the range of the upper or lower adjacent values) at more than 12 pulses, with a circle marker indicating its end points and using the same color for the study sites as in Figure 2. SAMT classified this trial as borderline accurate (see Figure S7). The seven trials flagged by SAMT as inaccurate were excluded from the statistics and are shown in red.

We calculated the 95% CIs of the fitted threshold parameters using the likelihood-ratio method, which had a small asymmetry, i.e., the threshold parameter typically was slightly closer to the lower bounds of the CIs (Figure S6). The widths of the CIs (Figure 4A) were large initially (some > 20% MSO) but rapidly decreased as more data were collected with each additional pulse, with the median CI width decreasing to less than 6% MSO at pulse number 10. By pulse number 25, the CI widths had a median within 3.6% MSO and were less than 6.2% MSO for all trials excluding the aforementioned outlier (6.8% MSO, Figure S7). Except during the first two pulses where a small but noticeable portion of the SAMT MTs were outside the CIs of the fitted thresholds, almost all the SAMT MTs fell within the CIs starting at pulse number 3, with only less than 1% of the trials (≤ 3) having MTs outside their respective CIs (Figure 4B). This further indicates that SAMT provided accurate MT estimates comparable to existing methods based on parametric models. Among the two trials with MT estimate outside the fitted threshold’s CIs near the trial end, one had MT estimates just within 0.2% MSO outside the corresponding CI. The other trial, however, consistently had large relative deviations (Figure S4) and was considered borderline accurate (8 responses out of the last 10 pulses), and could have been rejected by the TMS operator based on the response pattern, i.e., having 15 responses during the last 17 pulses (Figure S8).

**Figure 4:**
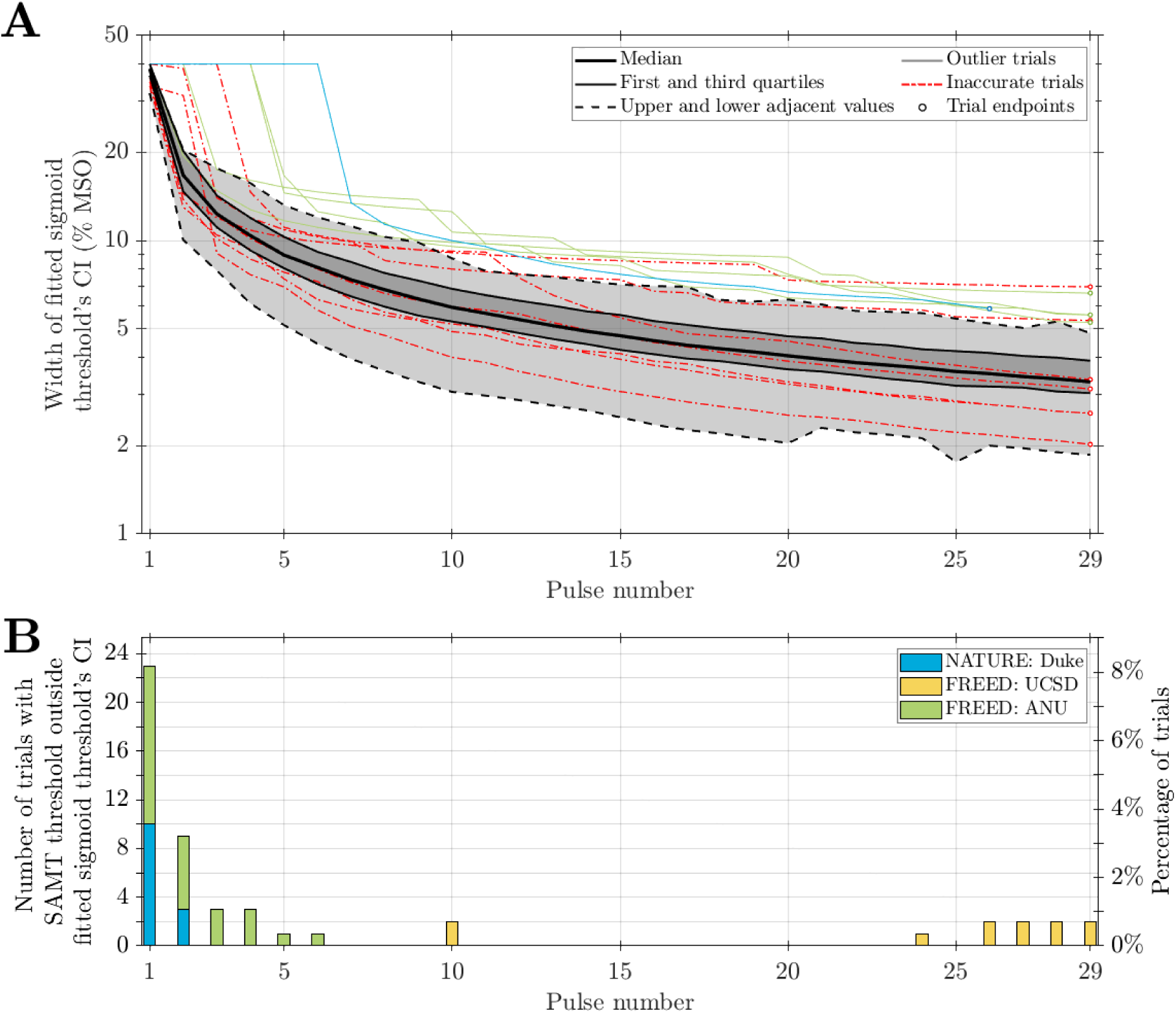
95% CI of fitted sigmoid threshold estimate and SAMT MTs outside CI. **A.** The widths of fitted thresholds’ CI as a function of pulse number. The CIs were wide for early pulses and likely inaccurate as the parameter fitting and CI calculation were not robust with a small number of data points. The thin lines show individual trials whose CIs were outliers for more than 12 pulses, following the same colors for the study sites as in Figure 2. **B.** The number of trials (left axis) and corresponding percentage of all trials (274, excluding inaccurate trials, right axis) for which the SAMT MTs were outside the CI at each pulse.

### Discussion and conclusions

In this study, we described the details of SAMT, a novel thresholding tool for TMS based on SA methods. The computational efficiency of the SA algorithm enables SAMT to run online without installation on a variety of platforms including not only computers, but also mobile phones and tablets. SAMT also implemented an improved version of the criteria proposed in the literature [13] for TMS operators to evaluate the accuracy of threshold estimates. This criteria identified trials with likely inaccurate MT estimates and instructed the user to rerun the procedures.

The thresholding data collected using SAMT in the clinical studies were used to assess the performance of the SA thresholding method for TMS in human participants. The results indicated that the convergence of SAMT MT to the fitted sigmoid threshold parameter was fast, and the deviations for most trials were small at the end of the procedure. With few exceptions, the deviations were within 1% MSO and 2% (269 out of 274 trials) and the MTs were within the 95% CIs of the fitted thresholds (272 out of 274 trials). Such deviations were also smaller than the variation observed within individual participants over time [32,38] as well as the pulse amplitude variability of TMS machines [44].

The deviations from the fitted thresholds and the latter’s CIs of the seven inaccurate estimation trials could be similarly calculated (Figures 2‒4 and S2‒S6, red markers or lines). Whereas five trials had large deviations that would be considered outliers above the upper adjacent value (black dashed line) for significant portions of the procedure (Figure 3), two had smaller deviations similar to those of trials considered accurate. The widths of their fitted thresholds’ CIs were similar to those of accurate trials (Figure 4). Therefore, given the limited number of pulses, the SA method alone could also result in inaccurate estimations similar to those previously demonstrated in parametric methods such as PEST; such trials, however, were rejected by SAMT’s detection of inaccurate estimation feature [13]. The detection method was simple and would potentially rely on the judgement of the TMS operator to deal with borderline cases in terms of the response count during the last 10 pulses (i.e., 2 or 8 responses) (Figures S7‒S8). To remove user judgment as a factor that may impact the accuracy of the final estimate, further refinement of the termination rule is needed to improve the classification of threshold estimate and develop automated termination rule. All inaccurate and almost all borderline accurate trials were from trials with fixed stepping, suggesting that adaptive stepping was more robust at obtaining accurate threshold estimates than fixed stepping.

Based on the convergence of MT deviations and CIs, flexible termination criteria could be developed that allow SAMT to adaptively stop the thresholding procedure sooner. Given the progression of threshold deviations, some trials achieved reasonable accuracy below 0.5% MSO between 15 to 25 pulses, and the procedure could have stopped before pulse number 25 if a robust termination rule were available. Accordingly, we have lowered the pulse number at which SAMT starts to assess threshold accuracy from 25 to 20 in the latest version (1.8.0). On the other hand, it may be possible to extend the procedure for those trials considered misestimation at the end of the trial (pulse number 29 for now) by a few steps to achieve accurate estimation without restarting the procedure. This case-specific extension may be especially appealing for the adaptive stepping method, where the convergence of the sequence does not necessarily slow down after a large number of steps. The statistics of MTs (Fig. 2A) obtained in this study could also serve as prior information to further accelerate convergence [45]. Another approach to improve convergence can be to optimize the initial step size depending on the starting pulse strength, since the optimal initial step depends on the slope of the sigmoidal input–output curve (2) at threshold [46], which is assumed to be correlated with the threshold [6,11].

The clinical studies also provided user feedback improving the functionality and user interface of SAMT. Since its use in the two clinical studies, SAMT has been updated and included many additional features such as visualization of the thresholding procedure, resulting in a simple, accurate, and useful thresholding tool with potential for wider adoption in the TMS community.

An important limitation of this work is that SAMT was used to estimate MTs in clinical studies whose primary objectives are not motor thresholding and which are therefore limited by the lack of ground truth threshold data. Obtaining experimental ground truth data is challenging since alternative MT estimation algorithms have their own accuracy limitations [6,13] and the added time for doing more MT titrations is not acceptable for most clinical studies. We therefore opted to compare SAMT’s MT estimates with a conventional method of modeling the response probability distribution with a sigmoid fitted *post-hoc* to the complete data from each SAMT sequence. While not providing a direct accuracy quantification, the reported deviation between the SAMT output and the fitted sigmoid threshold measures the agreement between the two methods. Thus, the presented experimental data in combination with the evidence from our prior computational study, which did include threshold ground truth, support the accuracy of SAMT.

In conclusion, we developed a new TMS motor thresholding tool, SAMT, and provided algorithm information, use instructions, and performance assessment in clinical studies, supporting SAMT’s practicality and accuracy.

## Availability statement

Latest versions of SAMT (currently 1.8.0) is available online at https://tms-samt.github.io [18]. The data that support the findings of this study are openly available at the Duke Research Data Repository with the following URL/DOI [47]: https://doi.org/10.7924/r44f2210t.

## Author contribution

AVP and SMG conceived, supervised, and secured funding and computational resources for the development of SAMT. BW and LMK wrote the online app and contributed concepts for preventing misestimation. BW maintained and updated SAMT, developed the confidence interval estimation methods, performed data analysis and visualization, and wrote the manuscript. VUS performed data analysis and contributed code to SAMT. ADN, JYC, NG, and YL (NATURE study) and ZJD, PBF, LGA, IH, YS, MP, HD, KR, ESG, NWB, JR, SB, and ATG (FREED study) incorporated SAMT into the respective study protocols, performed TMS thresholding using SAMT, and collected the SAMT data, with AND, ZJD, and PBF securing funding for the respective studies. All authors revised, commented on, and approved the final version of the manuscript.

## Data Availability

All data produced are available online at the Duke Research Data Repository with the following URL/DOI: https://doi.org/10.7924/r44f2210t

## Acknowledgments

Research reported in this publication is supported by the National Institutes of Health of the United States of America under Award Numbers R01 NS117405, RF1 MH124943, R01 MH129302, and R61 MH129471. LMK was supported by the European Union’s Horizon 2020 research and innovation program under Marie Sklodowska-Curie grant agreement No. 101027633. PBF is supported by a National Health and Medical Research Council of Australia Investigator grant (1193596). The content is solely the responsibility of the authors and does not necessarily represent the official views of the funding agencies.

The authors thank Dr. Marc A. Sommer and Neerav Goswami for testing and feedback on early versions of SAMT, David Chang Villacreses with the Duke Office for Translation and Commercialization for support with SAMT’s license, and Yiwen Zhang and Shivum Vaishnavi respectively for contribution related to the layout and visualization functions of SAMT. Preliminary results of this study were presented at the 2024 NYC Neuromodulation Conference (August 2024, New York, NY, USA) [48].

## Supplementary table

**Table S1.** Statistics of SAMT thresholds at end of trials.

|  |  | All studies | NATURE<br>Duke | FREED<br>Both sites | UCSD | ANU |
| --- | --- | --- | --- | --- | --- | --- |
| All SAMT trials |  | 281 | 65 | 216 | 117 | 99 |
| Misestimation trials |  | 7 | 2 | 5 | 3 | 2 |
| Sample size for statistics |  | 274 | 63 | 211 | 112 | 97 |
| SAMT MTs | Mean ± SD (%MSO) | 51.2 ± 10.1 | 51.4 ± 9.8 | 51.1 ± 10.2 | 49.9 ± 10.1 | 52.5 ± 10.2 |
|  | Median (%MSO) | 49.8 | 50.4 | 49.6 | 48.7 | 52.0 |
|  | Range (%MSO) | [22.3, 82.2] | [35.4, 82.2] | [22.3, 76.6] | [22.3, 70.7] | [33.6, 76.6] |
| | Normality test: $p$-value | 0.0491 | 0.798 | 0.0511 | 0.0577 | 0.751 |
|  | Number of outliers | 2 | 1 | 1 | 1 | 0 |
| Fitted sigmoid thresholds | Mean ± SD (%MSO) | 51.1 ± 10.1 | 51.4 ± 9.7 | 51.0 ± 10.3 | 49.8 ± 10.1 | 52.5 ± 10.3 |
|  | Median (%MSO) | 49.7 | 50.2 | 49.5 | 48.5 | 52.2 |
|  | Range (%MSO) | [23.3, 82.0] | [35.5, 82.0] | [23.3, 77.4] | [23.3, 70.6] | [33.6, 77.4] |
| | Normality test: $p$-value | 0.0391 | 0.899 | 0.0467 | 0.0528 | 0.759 |
|  | Number of outliers | 1 | 1 | 0 | 1 | 0 |
| SAMT MTs vs. fitted sigmoid thresholds | Equal variance test: F-statistics and $p$-value | 0.995, 0.965 | 1.009, 0.971 | 0.991, 0.947 | 1.005, 0.979 | 0.978, 0.912 |
| | Pearson correlation: $r$ and $p$-value | **0.9994, $\ll 0.01$** | **0.9995, $\ll 0.01$** | **0.9994, $\ll 0.01$** | **0.9994, $\ll 0.01$** | **0.9995, $\ll 0.01$** |
| | Intraclass correlation: ICC and $p$-value | **0.9994, $\ll 0.01$** | **0.9995, $\ll 0.01$** | **0.9994, $\ll 0.01$** | **0.9993, $\ll 0.01$** | **0.9995, $\ll 0.01$** |
| Absolute deviations | Mean ± SD (%MSO) | 0.06 ± 0.35 | 0.02 ± 0.31 | 0.07 ± 0.36 | 0.11 ± 0.36 | 0.01 ± 0.35 |
|  | Median (%MSO) | 0.06 | 0.05 | 0.06 | 0.09 | 0.04 |
|  | Range (%MSO) | [-2.14, 1.25] | [-1.20, 0.64] | [-2.14, 1.25] | [-1.35, 1.25] | [-2.14, 0.66] |
| | Normality test: $p$-value | **0.005** | 0.689 | **0.008** | 0.097 | 0.039 |
|  | Number of outliers | 12 | 1 | 11 | 6 | 6 |
| | $t$-test: $p$-value | **0.0098** | 0.628 | **0.009** | **0.002** | 0.724 |
| | Correlation with threshold: $r$ and $p$-value | -0.060, 0.320 | 0.163, 0.202 | -0.113, 0.101 | 0.083, 0.381 | **-0.317, 0.002** |
| Relative deviations | Mean ± SD | 0.11% ± 0.77% | 0.03% ± 0.64% | 0.14% ± 0.81% | 0.21% ± 0.93% | 0.06% ± 0.63% |
|  | Median | 0.12% | 0.09% | 0.12% | 0.17% | 0.07% |
|  | Range | [-4.86%, 2.64%] | [-2.47%, 1.35%] | [-4.86%, 2.64%] | [-4.86%, 2.64%] | [-2.90%, 1.69%] |
| | Normality test: $p$-value | **0.002** | 0.280 | **0.002** | 0.011 | 0.208 |
|  | Number of outliers | 15 | 4 | 10 | 7 | 7 |
| | $t$-test: $p$-value | 0.015 | 0.754 | 0.012 | 0.019 | 0.333 |
| | Correlation with threshold: $r$ and $p$-value | -0.018, 0.771 | 0.124, 0.333 | -0.049, 0.494 | 0.103, 0.277 | **-0.283, 0.005** |

Notes: (1) Entries that are statistically significant at a level of 0.01 are highlighted in **bold font**. (2) Outliers are defined as being outside the upper or lower adjacent values, which are 1.5 times the interquartile range above or below the third and first quartiles, respectively. (2) Any $p$ -values reported as $\ll 0.01$ indicate extremely small numbers calculated by MATLAB that are likely not reliably accurate, e.g., $10^{-88}$ or even 0, with the latter likely due to calculated $p$ -values being smaller than the limit of double-precision floating numbers of $2.225 \times 10^{-308}$ in MATLAB.

## Supplementary figures

**Figure S1:**
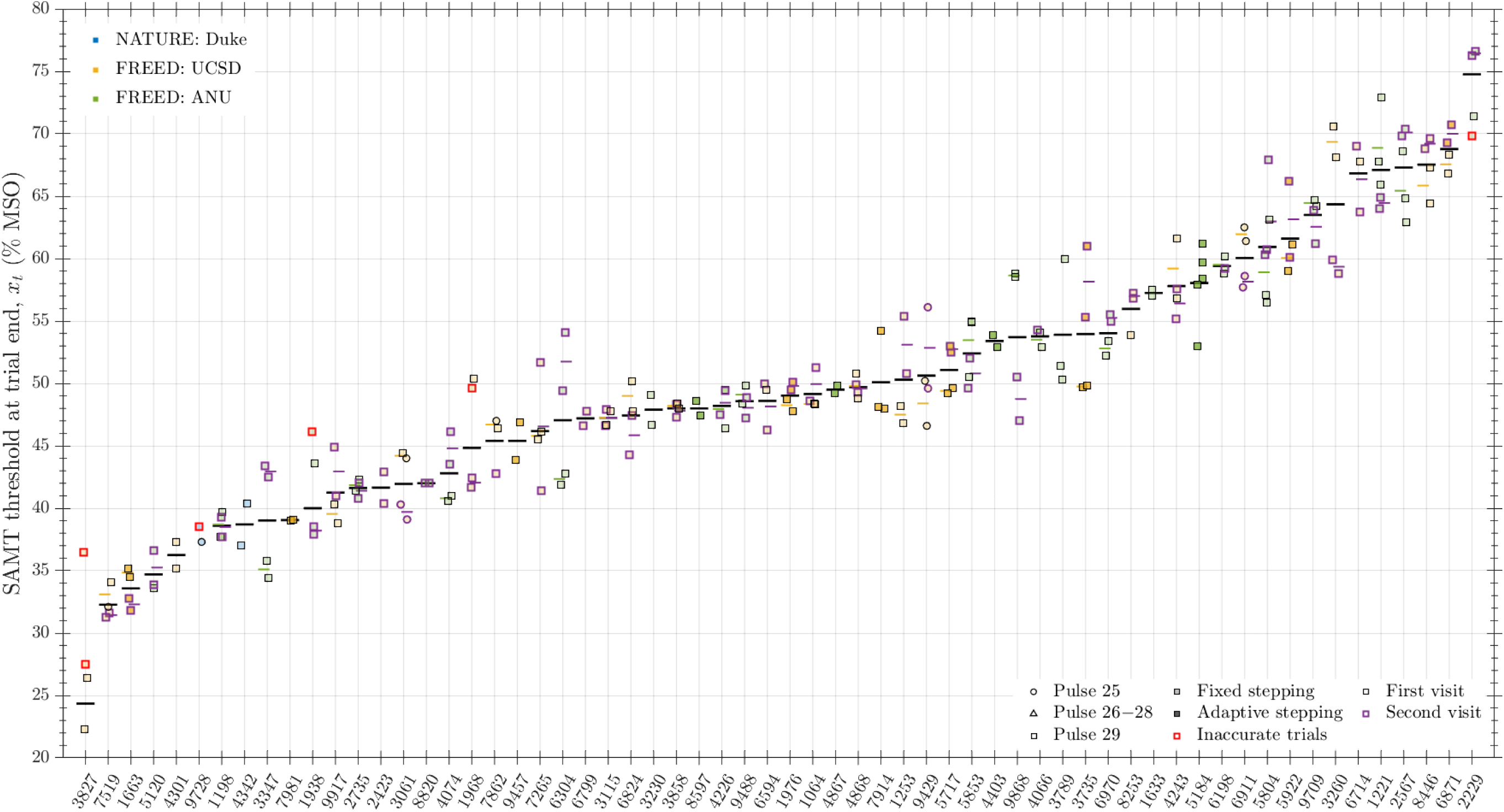
Repeated measurements for individual participants, ordered by average threshold. Marker shape and shading color follow the same as in Figure 2. Purple outlines indicate measurements on a second visit post-treatment in the FREED study. Thick horizontal lines in black indicate the mean, excluding inaccurate trials (red outline), and are not plotted if there was only single accurate data point (participant 9728). For FREED study participants, if there were multiple trials on either the first or second visit, the averages of the visits are shown with short thin lines in the group color (yellow for UCSD, and green for ANU) and purple, respectively. Data available only for second visit for participants 2423, 8820, and 6799. Note: random 4-digit numbers were assigned to the participants for reporting here.

**Figure S2:**
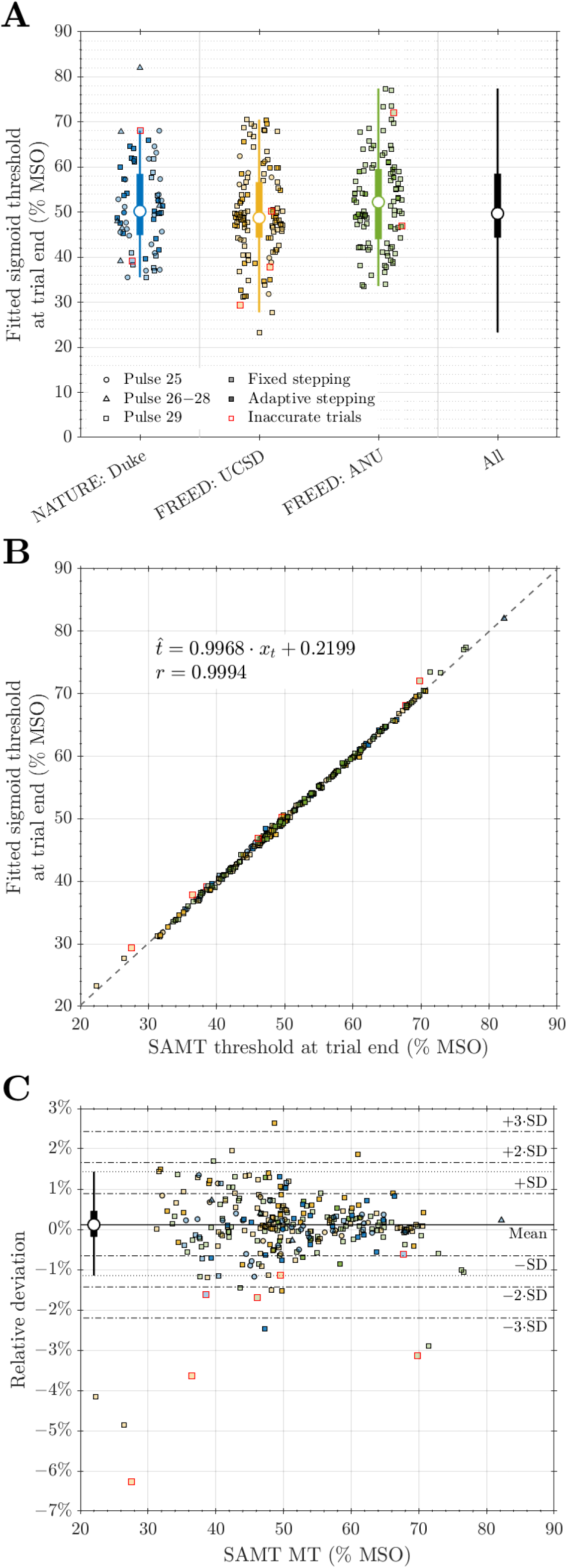
Fitted sigmoid threshold parameter and relationship with SAMT MT at trial end. **A.** Distribution of fitted threshold parameters at the end of the trial, in similar format as Figure 2A. **B.** Scatter plot demonstrating high correlation between fitted thresholds and SAMT MTs. **C.** A modified Bland-Altman plot shows distribution of relative deviations at the trial end versus the SAMT threshold estimates, in similar format as Figure 2B.

**Figure S3:**
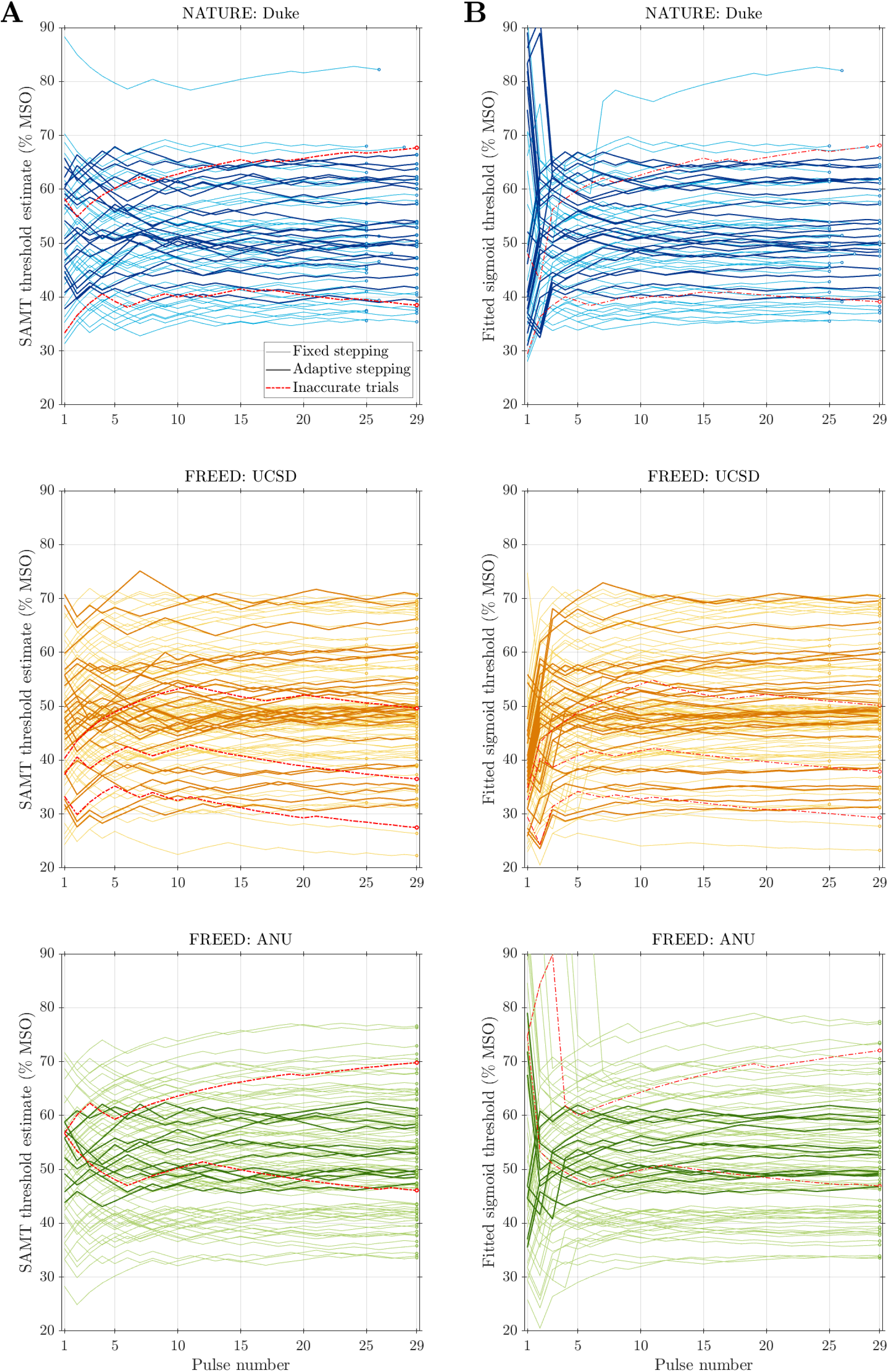
Progression of SAMT MT and fitted sigmoid threshold parameter. SAMT MTs (**A**) and fitted threshold parameter (**B**) as a function of pulse number for the three study sites. Trial with fixed and adaptive stepping have light and dark line colors, respectively, and the trial end point is marked with a circle. The back-and-forth stepping around the putative threshold is evident for most individual trials as the procedure converges. In contrast, the inaccurate trials have one-sided stepping, especially towards the end of the procedure, which does not converge on a specific stimulation amplitude.

**Figure S4:**
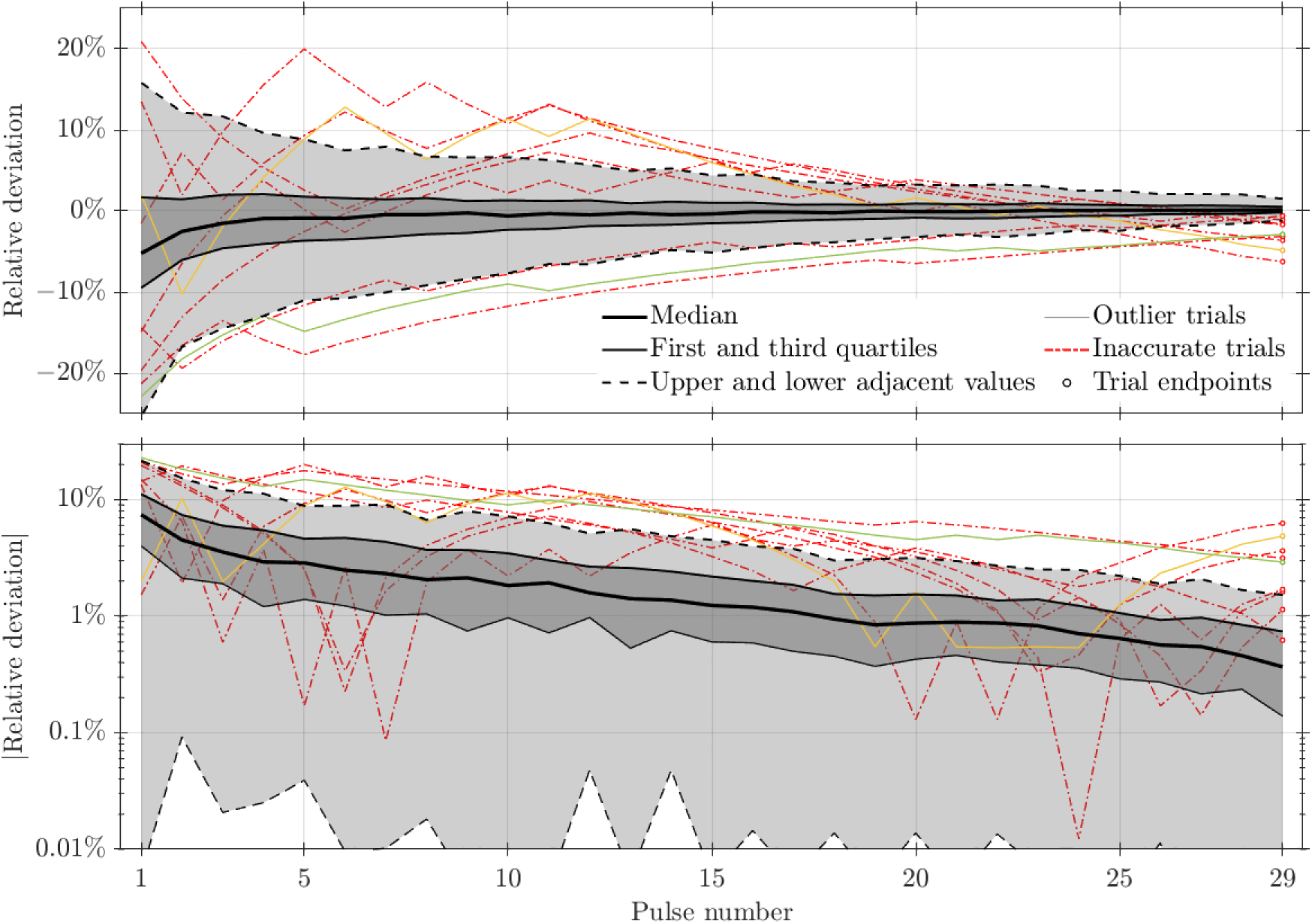
Relative deviation of SAMT MTs versus number of pulses. Shown similarly as the absolute deviation in Figure 3. Two individual trials consistently had large deviations considered as outliers at more than 12 pulses. One trial (green lines) was also an outlier trial for absolute deviations (Figures 3 and S7) and the other one (yellow lines) had MTs outside the fitted parameter’s CI towards the end of the trial (Figures 4B and S8). Both were both considered borderline accurate by SAMT.

**Figure S5:**
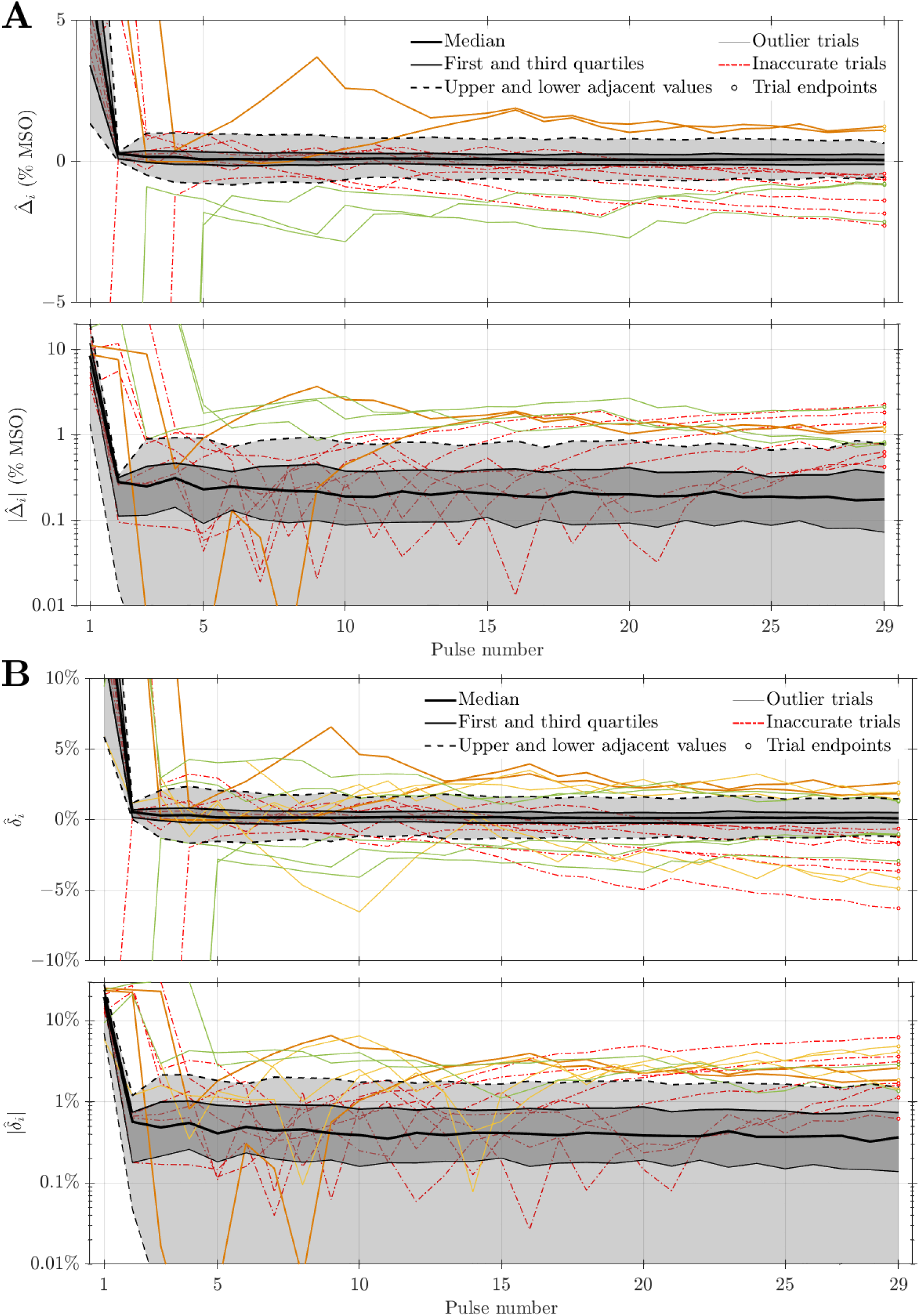
Pulse-by-pulse deviation of SAMT MTs. Shown similarly as the *post-hoc* deviations in Figures 3 and S4. Unlike the *post-hoc* deviation, which used the fitted sigmoid threshold parameter obtained from the entire procedure as reference, the pulse-by-pulse deviations were calculated against the sigmoid thresholds fitted with partial data up to the same pulse number. Due to the tighter distributions, individual trials are shown only if they have deviations considered as outliers at 18 or more pulses. **A.** Absolute deviation Δ̂_*i*_. **B.** Relative deviation *δ̂*_*i*_.

**Figure S6:**
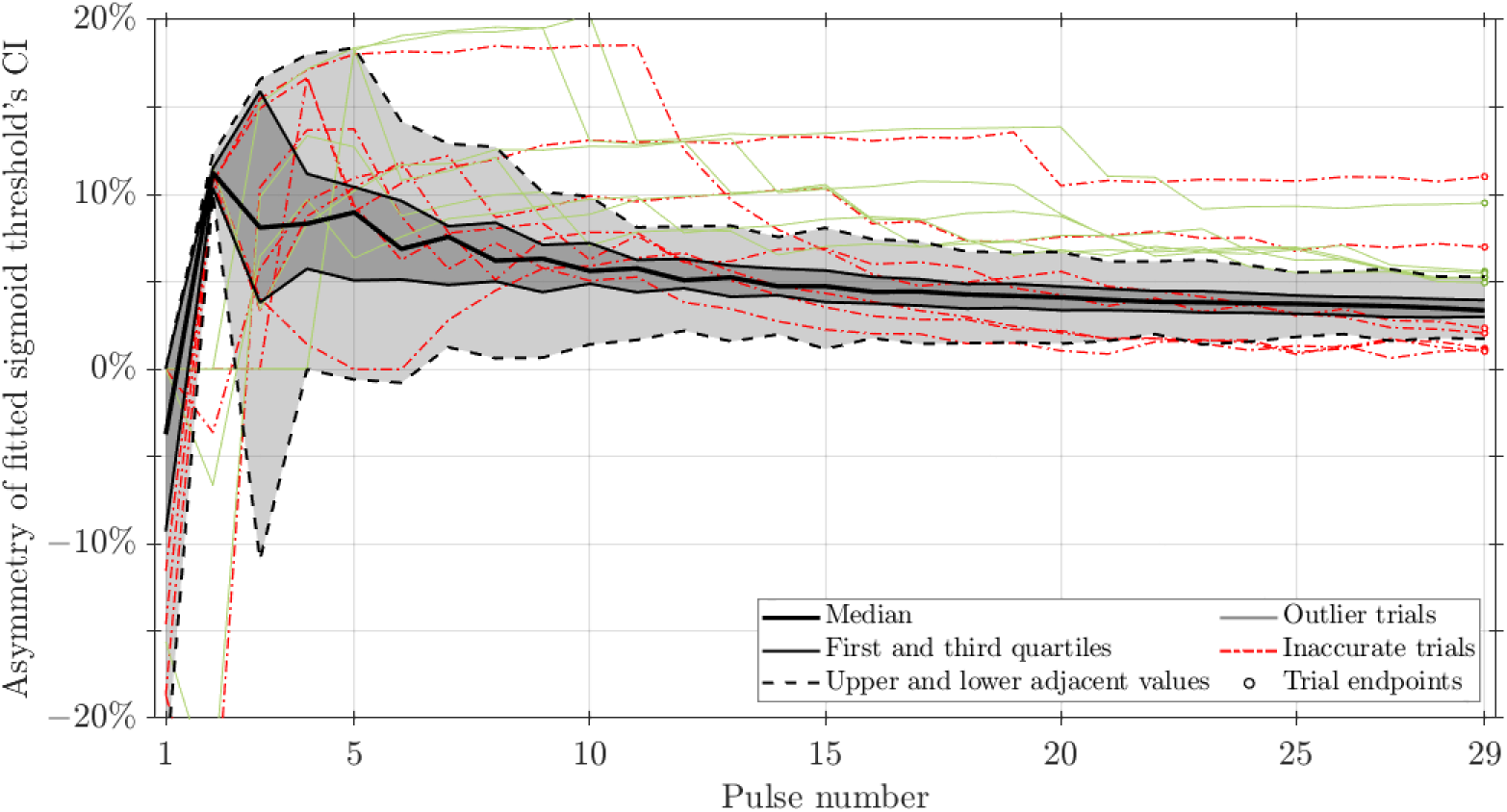
Asymmetry of the fitted sigmoid thresholds’ 95% confidence intervals (CI). The CIs of fitted sigmoid thresholds were slightly asymmetric, with the asymmetry here defined as the difference between the widths of the upper (Δ*U*_*i*_ = *U*_*i*_ − *t̂*_*i*_) and lower (Δ*L*_*i*_ = *t̂*_*i*_ − *L*_*i*_) intervals of the CI as a percentage of the total CI width: (Δ*U*_*i*_ − Δ*L*_*i*_)/(*U*_*i*_ − *L*_*i*_), where the upper and lower intervals were between the upper bound *U*_*i*_ and the parameter *t̂*_*i*_, and between the parameter and the lower bound *L*_*i*_, respectively. This metric was equivalent to the percentage difference between the width of the upper interval and half the total width, if the CI had been positioned symmetrically with the parameter at the midpoint. Here, the overall positive asymmetry (except at the very beginning) indicates that the upper intervals were slightly longer, and the threshold parameters were located in the lower halves of the CIs.

**Figure S7:**
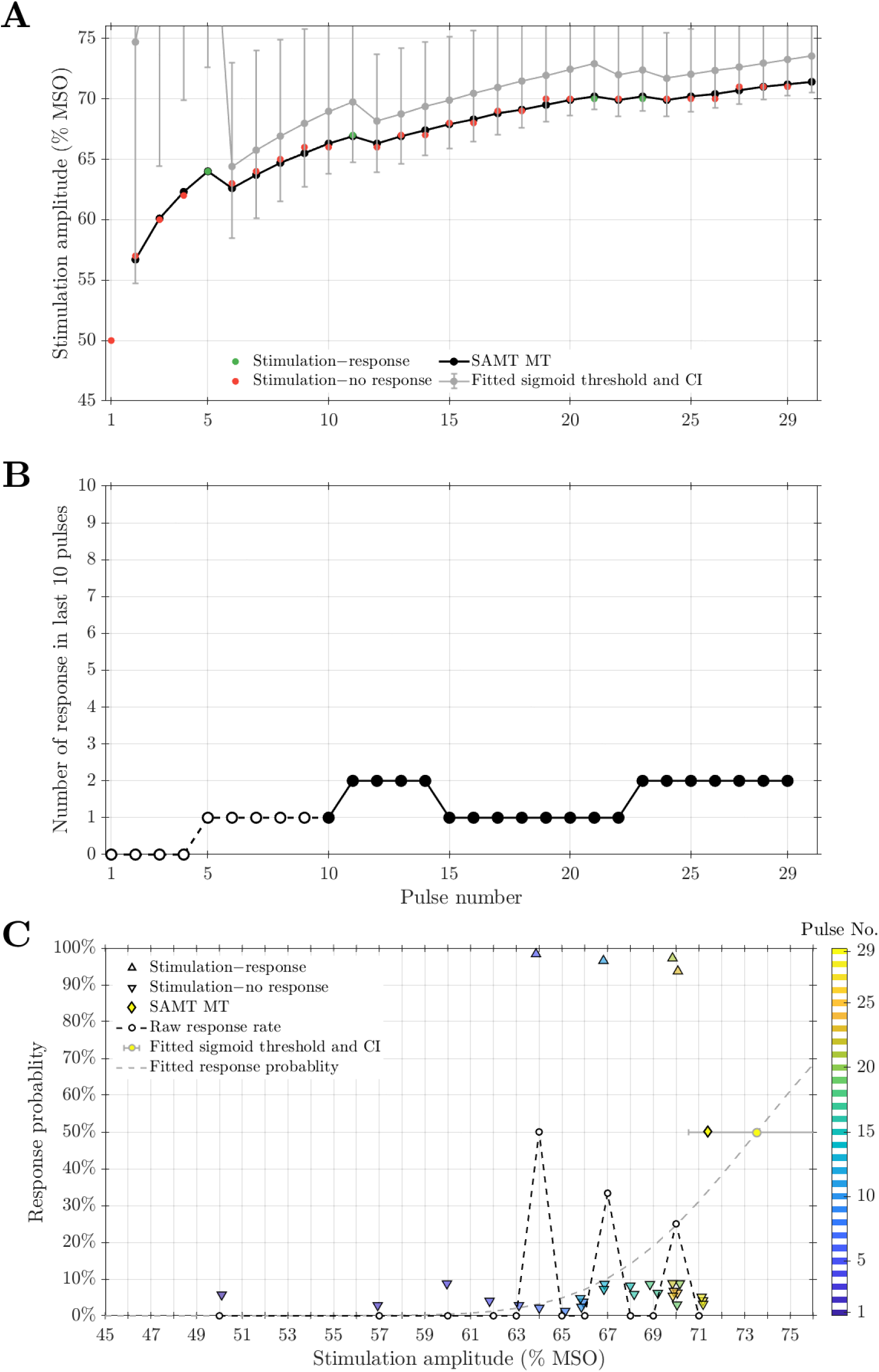
An example of a borderline accurate trial with few responses during the last 10 pulses. **A.** The stimulation amplitude and whether a response was evoked (green) or not (red) as the trial progressed. The SAMT MTs (black) for each step are plotted at the next step, as they determined the pulse amplitude to be applied (after rounding). The fitted sigmoid thresholds and their 95% CIs (gray markers and error bars) are similarly shifted to align with the corresponding SAMT MTs. The fitted thresholds were consistently larger than the SAMT MTs and had wide CIs. **B.** The response count during the last 10 pulses. Before pulse number 10, the count was for less than 10 pulses as indicated by open markers and dashed line. **C.** Summary information at the end of the trial. For each stimulation pulse, a response is shown as an upper triangle at near 100% response rate and a no-response is shown as a lower triangle near 0% response rate, with the color indicating the pulse number. The markers are slightly offset for clarity. The dashed line and circle markers show the raw response rate for each amplitude that was used during the trial. The SAMT MT estimate is shown as a diamond at 50% response rate. The fitted threshold and CI are given by the gray marker and horizontal error bars, respectively, and the response probability of the underlying model is given by the gray dashed line. The SAMT MT was larger than any stimulation pulse amplitude during the trial and had a large deviation compared to the fitted threshold.

**Figure S8:**
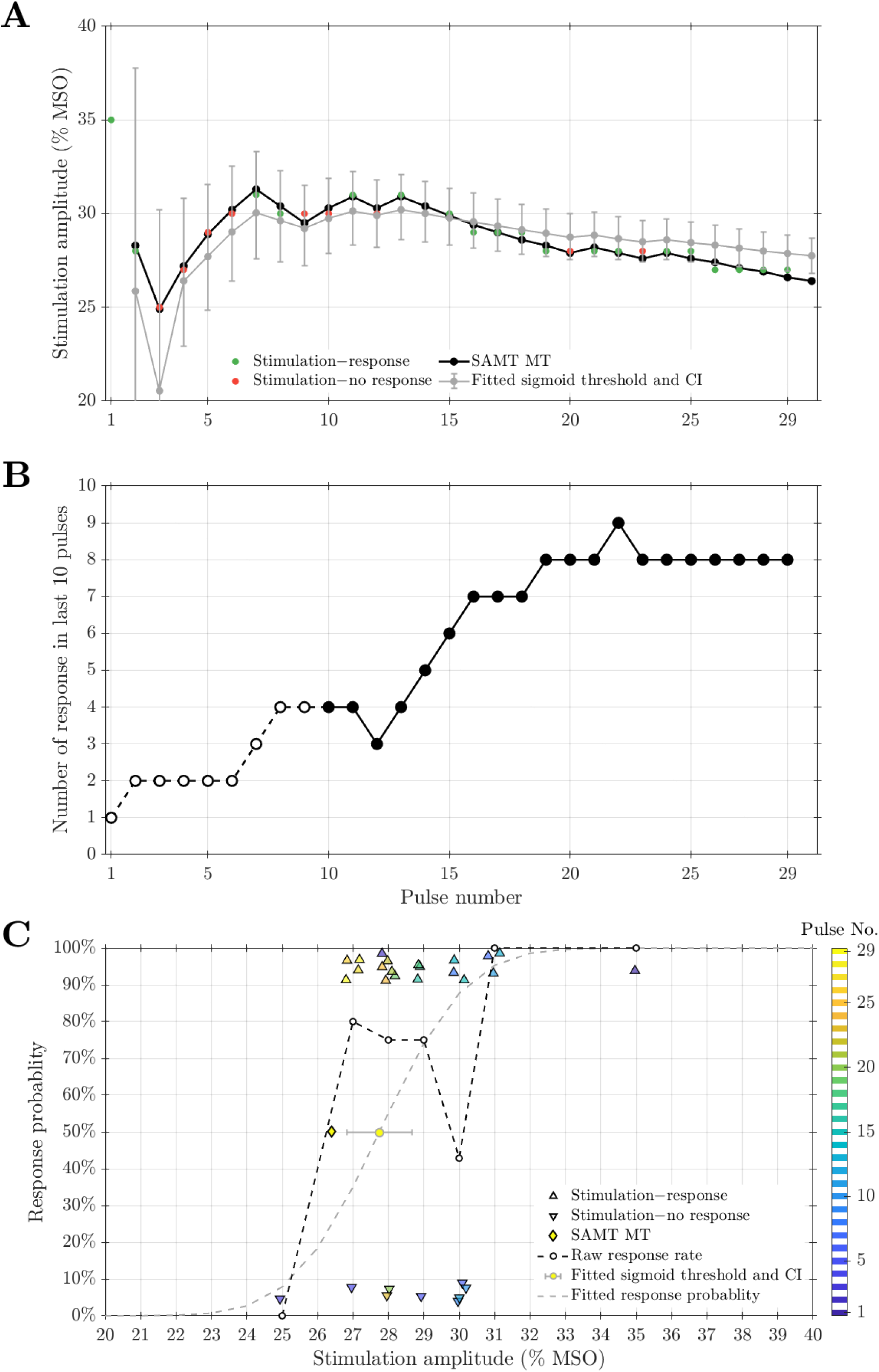
An example of a borderline accurate trial with many responses during the last 10 pulses. Same format as Figure S7. The SAMT MT drifted outside the fitted sigmoid thresholds’ 95% CIs starting at pulse number 26 and, at the last pulse, was smaller than all but one stimulation amplitudes (pulse number 2) and quite far outside the fitted threshold’s CI.

## Footnotes

1 Also known as MLE1 [6], adaptive threshold hunting (ATH) or estimation [11], best-PEST [11,12], or ML-PEST [13], and implemented in MTAT [14] and ATH-tool [15].

2 And DCS-H was deprecated in version 1.7.5.

3 In this context and the remainder of the paper, “trials” refers to MT titrations, and not clinical trials.

4 For this study, the minimum number of pulses was conservatively selected as 25 pulses initially—corresponding to a median relative error of 1.3% in the modeling study—and used up to SAMT version 1.7.5. Based on preliminary results, the threshold was lowered to 20 pulses in version 1.8.0.

